# Incidence and severity of SARS-CoV-2 infections in people with primary ciliary dyskinesia

**DOI:** 10.1101/2022.10.14.22281075

**Authors:** Eva SL Pedersen, Leonie D Schreck, Myrofora Goutaki, Sara Bellu, Fiona Copeland, Jane S Lucas, Marcel Zwahlen, COVID-PCD patient advisory group, Claudia E Kuehni

## Abstract

**Importance:** Early in the COVID-19 pandemic, chronic respiratory disease was considered a risk factor for severe COVID-19 disease. Studies have confirmed a higher risk of intensive care unit admission and mortality in people with chronic pulmonary obstructive disease and cystic fibrosis, but there is little data in people with primary ciliary dyskinesia (PCD).

**Objective:** To study incidence of SARS-CoV-2 and its risk factors in people with PCD from May 2020 to May 2022. We also describe the severity of COVID-19 symptoms in this population and factors associated with severity.

**Design, setting, and participants:** We used data from COVID-PCD, an international participatory cohort study following people with PCD through the COVID-19 pandemic. The study is based on self-reported weekly online questionnaires, available in five languages, adapted to children, adolescents, and adults. COVID-PCD invites people with PCD of any age to participate.

**Exposures:** SARS-CoV-2

**Main Outcomes:** Incidence of reported positive test of SARS-CoV-2 and reported severity of symptoms.

**Results:** By May 2022, 728 people with PCD participated (40% male, median age 27 years; range 0-85). The median weeks of follow-up was 60 (range 1-100). Eighty-seven (12%) reported a SARS-CoV-2 infection at baseline or during follow-up and 62 people reported an incident SARS-CoV-2 infection during 716 person-years of follow-up (incidence rate 9 per 100 person years; 95%CI 7-11). Using Poisson regression, we found that age above 14 years was associated with lower risk of infection (IRR 0.42, 95%CI 0.21-0.85) but the strongest predictors were exposure to Delta (IRR 4.52, 95%CI 1.92-10.6) and Omicron variants (IRR 13.3, 95%CI 5.2-33.8) compared to the original strain. Severity of disease was mainly mild; 12 (14%) were asymptomatic and 75 (86%) had symptoms among whom 4 were hospitalized. None needed intensive care and nobody died. Using Poisson regression, we found that comorbidity (IRR 1.93, 95%CI 1.40-2.64) and being infected during the period when the Delta variant was predominant (IRR 1.43, 95%CI 1.07-1.92) were associated with more reported symptoms.

**Conclusion and Relevance:** People with PCD do not seem to have a higher incidence of SARS-CoV-2 infections nor higher risk of severe COVID-19 disease than people from the general population.

**Key Points:** *Question:* What is the incidence and severity of COVID-19 in people with primary ciliary dyskinesia and which factors are associated with reporting a SARS-CoV-2 infection and risk of severe disease?

*Findings:* This international cohort of 728 people with primary ciliary dyskinesia followed for two years during the COVID-19 pandemic found a low incidence of reported SARS-CoV-2 in people with primary ciliary dyskinesia and mainly mild disease. The strongest predictor of incidence and severity was virus variant.

*Meaning:* People with PCD do not seem to have a higher incidence of SARS-CoV-2 infections nor higher risk of severe COVID-19 disease than people from the general population.

## Introduction

Early in the COVID-19 pandemic, chronic respiratory disease was considered a risk factor for severe COVID-19, the disease caused by severe acute respiratory syndrome coronavirus 2 (SARS-CoV-2) ^1-3^. Later studies confirmed a higher risk of intensive care unit admission and mortality in people with chronic pulmonary obstructive disease and cystic fibrosis (CF) ^4-6^. Asthma, in contrast, does not seem to increase the risk for severe COVID-19 ^7^. The situation remains unclear for people with rarer respiratory diseases, such as Primary Ciliary Dyskinesia (PCD). PCD is a genetic disease with a prevalence of approximately 1 in 10,000 or less ^8,9^. It is characterized by mutations that impair the function of motile cilia leading to chronic upper and lower airway disease and lung function impairment early in life ^10-16^. To gather information about COVID-19 in people with PCD, in May 2020 we set up the COVID-PCD, an online international participatory cohort study to follow people with PCD during the COVID-19 pandemic ^17^. In March 2021, we published preliminary results which showed a low incidence of 3.2 cases per 100 person-years (PY) and mainly mild disease among 24 people infected with SARS-CoV-2 ^18^. Since then we have acquired more data, vaccinations against COVID-19 became available, national advice for isolation changed, and different virus variants of concern appeared, all of which influenced incidence and severity of SARS-CoV-2 infections in healthy and diseased populations ^6,19,20^.

This paper describes the incidence of SARS-CoV-2 and its risk factors in people with PCD from May 2020 to May 2022. We also describe the severity of COVID-19 disease in this population and factors associated with severity.

## Methods

### Study design

We used data from COVID-PCD, an international participatory cohort study following people with PCD through the COVID-19 pandemic (clinicaltrials.gov: NCT04602481). COVID-PCD was initiated by people with PCD who wished to gather evidence on risk of COVID-19 in people with PCD. It is a participatory study where representatives from PCD patient support groups all over the world had a major role in developing study logistics, recruitment of study participants, and dissemination of results. COVID-PCD invites people with PCD of any age to participate. Questionnaires, based on FOLLOW-PCD ^21^, were adapted to three age-groups; children aged 0-13 years (completed by parents), adolescents aged 14-17, and adults aged 18 years and older. Questionnaires can be completed in five languages (English, German, Spanish, Italian, and French). Recruitment started on May 30^th^, 2020. All participants aged 14 years and older and parents of children below 17 years gave online consent. The ethics committee of the canton of Bern, Switzerland, approved the study (Study ID: 2020-00830). This manuscript was prepared following the STROBE guidelines ^22^.

### Study procedures and questionnaires

Study procedures have been published ^17^. In short, the study was advertised through PCD support groups in the UK, in North America, Switzerland, Germany, Australia, Italy, Spain, and France. Participants registered to participate through the COVID-PCD study website (www.covid19pcd.ispm.ch) and then received a link via email to the baseline questionnaire which included questions on SARS-CoV-2 infections experienced prior to study entry, information about PCD diagnosis, clinical history, current symptoms, environment, and lifestyle. Participants then received weekly follow-up questionnaires with questions on incident SARS-CoV-2 infections and symptoms. In December 2021 after 80 weeks of follow-up, we reduced the frequency of follow-up questionnaires to once every four weeks. Participants also received two extra questionnaires which studied specific questions in detail; one focused on use of face masks and one on COVID-19 vaccinations ^23,24^. The present paper includes all study participants who completed the baseline questionnaire.

### Definitions of outcomes and exposures

We defined a case as a reported positive test of SARS-CoV-2, including antigen, polymerase chain reaction (PCR), and antibody tests, reported at baseline or during follow-up. An incident case was defined as a positive test for SARS-CoV-2 reported in a follow-up questionnaire in people who had not reported an infection in the baseline-questionnaire. We defined follow-up time as days between date of study entry and date of last completed questionnaire and calculated person-years (PY) as follow-up time/365.25.

We studied whether the following factors were associated with higher risk of reporting a SARS-CoV-2 infection based on findings from previous studies: age, sex, and country ^25,26^, vaccination status ^27,28^, and virus variant ^29^. We defined participants as vaccinated if they reported at least one dose of vaccine in the baseline, in a follow-up, or the special questionnaire on COVID-19 vaccinations. Date of full vaccine protection was defined as 14 days after receiving the second dose of Pfizer/BioNTech, Moderna, AstraZeneca, or Sinovac, or as 14 days after receiving first dose of Janssen, Johnson and Johnson. In participants who only reported the first dose (n=38), we defined date of full vaccine protection as 44 days (1.5 months) after, assuming these people also got vaccinated a second time but did not report this date. In participants who only reported the date of the booster dose (n=3), we defined date of full vaccine protection as 135 days (3.5 months) before. This interval was chosen as the booster is recommended 4 months after the second dose, and full protection is assumed to occur 14 days after the second dose ^30^. We did not have information about virus variant for each infected person, and we therefore as proxy defined virus variant based on reported time of infection. For each of the major regions: UK, Europe other than UK, North America, Australia, and other regions, we divided time into periods in which one virus variant caused more than 50% of all cases based on OurWorldInData ^31^. Apart from the original strain, we divided time into periods based on the three main variants of concern: Alpha, Delta, and Omicron, which were the only variants that caused more than 50% of all cases in a larger area. For simplification, we chose the closest first day in a month for the change in two periods, so that for example for the UK, the period dominated by the original strain went from January 1, 2020, to December 31, 2020, while the period dominated by the Alpha variant went from January 1, 2021, to May 31, 2021. The definition of each period for the major regions are described in detail in the supplementary information (supplementary table 1).

Severity of COVID-19 disease was defined based on self-reported data categorised as asymptomatic (no symptoms), mild (light fever, mild cough), and moderate (high fever and other symptoms such as headache and cough). We also assessed hospitalization due to COVID-19 and number of days in hospital. We studied whether the following factors were associated with self-reported severity and number of reported symptoms based on previous literature ^4,32-34^: age, sex, self-reported bronchiectasis at baseline, and self-reported FEV_1_ as proxy for severity of PCD, comorbidities, and COVID-19 vaccination. We dichotomised reported FEV_1_ as above and below 60% predicted. There is no suggested cut-off in people with PCD, however, in people with CF, an FEV_1_ below 60% has been associated with severe disease ^35^ and lung function impairment is comparable in people with PCD and CF ^36^. Due to sample size restrictions, we could not include single comorbidities as risk factors and therefore created a composite variable defined as any of the following: hypertension, diabetes, heart disease or heart failure, cancer, Crohn’s disease, stroke, and BMI above 30. The formulation of questions to assess SARS-CoV-2, bronchiectasis, FEV_1_, and comorbidities are reported in supplementary table 2.

### Statistical analyses

We describe the proportion of people with PCD who reported an infection with SARS-CoV-2 as number of total SARS-CoV-2 infections reported at baseline or during follow-up divided by total number of included participants. We describe incidence of reported SARS-CoV-2 as number of incident cases during follow-up per 100 PY. We studied predictors of reporting a SARS-CoV-2 infection using multivariable Poisson regression with a logarithmic link function and follow-up interval in days as offset variable and report results as incidence rate ratios (IRR). We included age stratified in groups, sex, country, vaccination status, and virus variant as predictors. Follow-up intervals for each individual correspond to the number of completed follow-up questionnaires and the interval length was days between completion of one follow-up questionnaire until completion of the next follow-up questionnaire. For each follow-up interval, reported infection with SARS-CoV-2 was defined as ‘No’ or ‘Yes’ corresponding to whether the person reported an infection during the follow-up interval. For our main analysis, only first infections with SARS-CoV-2 were included as the outcome because only 8 people reported a second infection. As a sensitivity analysis, we also included second infections as outcome. For participants without vaccination status information, we set full vaccination status to “no”, as all participants with no vaccination information completed the last follow-up questionnaire before February 1, 2021, when we had not yet asked about vaccination status in the follow-up questionnaires and very few participants had been vaccinated. We included age groups in the final Poisson regression after exploring linear and non-linear effects of age as a continuous variable. To study factors associated with severity of COVID-19, we tested whether age, sex, bronchiectasis, FEV_1_, comorbidity, and vaccination status differed by self-reported severity using χ2. We also created a variable representing the number of reported symptoms by adding together all reported symptoms for each individual resulting in a count-variable from 0-16. To study associated factors, we used multivariable Poisson regression with number of symptoms as outcome and age groups, comorbidities, virus variant, and vaccination status as predictors. We used STATA version 15 for all statistical analysis.

## Results

### Study population

We included 728 people with PCD with a median age of 27 years (range 0-85) of whom 434 (60%) were female (table 1). Follow-up data were available for 664 participants (90%). The median number of weeks of follow-up was 60 (range 1-100 weeks, interquartile range 26-88) and on average, participants completed 57% of the questionnaires (standard deviation 30%). One person died of reasons unrelated to SARS-CoV-2 during follow-up, and another nine people left the study (one received an alternative diagnosis than PCD and eight did not give a reason). Participants came from 48 countries with most from the UK (20%), USA (18%), and Germany (14%). At baseline, 453 (62%) reported bronchiectasis and of the 435 (60%) who reported recent lung function test results, 226 (52%) reported FEV1 below 60% predicted. The most common comorbidity was asthma reported by 25%. Most adolescents and adults were vaccinated against COVID-19; 88% of the 15-49-year-olds and 95% of those aged 50 years and older.

**Table 1:** Characteristics of people with primary ciliary dyskinesia included in COVID-PCD study (N=728)

| Characteristics | Total, N=728<br>N(%) |
| --- | --- |
| **Age categories** |  |
| 0-14 years | 228 (31) |
| 15-49 years | 381 (52) |
| 50 years and above | 119 (16) |
| **Sex** |  |
| Male | 292 (40) |
| Female | 434 (60) |
| Other | 2 (0) |
| **Country** |  |
| United Kingdom | 145 (20) |
| USA | 128 (18) |
| Germany | 103 (14) |
| Switzerland | 48 (7) |
| Italy | 47 (7) |
| France | 42 (6) |
| Australia | 32 (4) |
| Other European countries | 123 (17) |
| Other non-European countries | 59 (8) |
| **Severity of PCD** |  |
| Congenital heart disease | 53 (7) |
| Bronchiectasis | 453 (62) |
| FEV1 percent predicted (n=435) |  |
| >60% predicted | 209 (48) |
| <60% predicted | 226 (52) |
| **Comorbidity (at baseline)** |  |
| Hypertension* | 38 (8) |
| Diabetes* | 13 (3) |
| Heart disease or heart failure* | 11 (2) |
| Cancer* | 6 (1) |
| Crohn's disease or Colitis ulcerosa* | 12 (3) |
| Stroke ever* | 3 (1) |
| BMI>30* | 53 (11) |
| Any comorbidity* | 98 (21) |
| **Fully vaccinated against COVID-19**<br>**(N=569#)** |  |
| 0-14 year-olds | 79 (45) |
| 15-49 year-olds | 250 (88) |
| 50 year or older | 102 (94) |

\*only asked in adults aged 18 years or older, N=477 #Calculated among those who completed a special questionnaire on vaccines or completed vaccination information in a follow-up questionnaire.

### Incidence of SARS-CoV-2 infections

In total, 87 participants (12%) reported a SARS-CoV-2 infection at baseline or during follow-up (supplementary table 3), and 8 participants reported a second SARS-CoV-2 infection during follow-up. The number of infections differed between months (figure 1). Most infections were reported during the time when Delta and Omicron were dominant. From March 2020 to May 2021, most infections were reported for adults while from May 2021 to May 2022, most infections were reported for children (data not shown). During 716 PY of follow-up, 62 participants reported a SARS-CoV-2 infection corresponding to a cumulative incidence rate of 8.7 per 100 PY (95% confidence interval (CI) 6.8-11.0) (supplementary table 3). In the multivariable Poisson model, we found that age above 14 years was associated with lower risk of reported infection (IRR 15-49 years 0.42, 95%CI 0.21-0.85) (table 2). Compared to the UK, people living in other countries were less likely to report an infection (e.g. IRR Germany 0.46, 95%CI 0.21-1.03). The strongest predictors of a SARS-CoV-2 infection were exposure to Delta (IRR 4.52, 95%CI 1.92-10.6) and Omicron variants (IRR 13.3, 95%CI 5.2-33.8).

**Table 2:**
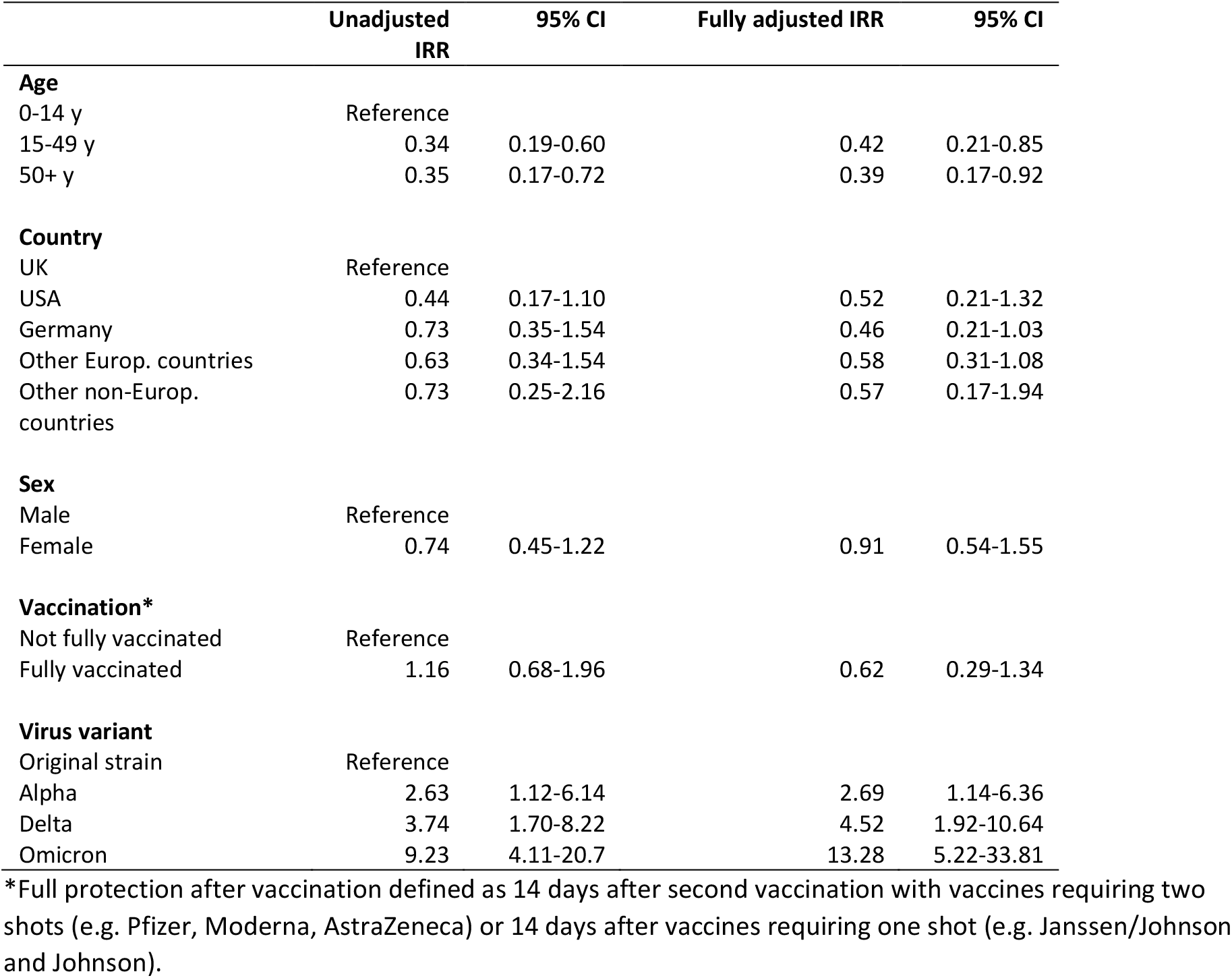
Incidence rate ratio (IRR) of SARS-CoV-2 from a poisson regression analysis including age, country of residence, sex, vaccination status, and virus variant among people with PCD included in the COVID-PCD study (n=62).

**Figure 1:**
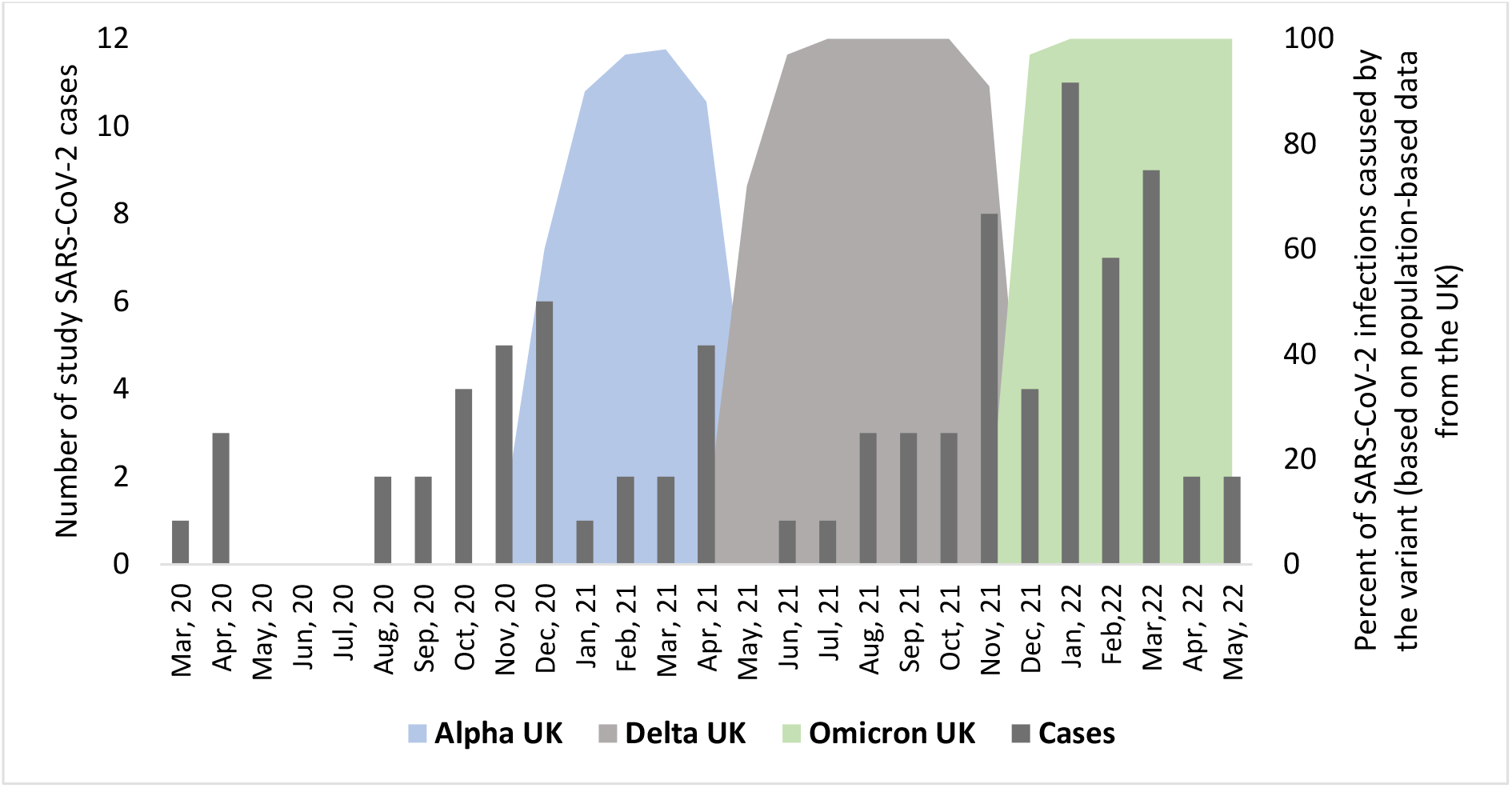
Timing of infections among people with primary ciliary dyskinesia from March 2020 to April 2022 (n=87), excluding re-infections, and information about dominant SARS-CoV-2 virus variants.

### Severity of SARS-CoV-2 infections

Severity of disease was overall mild in the 87 participants infected with SARS-CoV-2. Twelve participants (14%) reported an asymptomatic infection, 50 (57%) reported mild symptoms, and 25 (29%) reported moderate symptoms. Only four people reported hospitalization due to COVID-19. Of these, 2 reported mild symptoms while the other two reported moderate symptoms. The longest hospital stay was 9 days, and nobody was treated in the intensive care unit or died. We found little difference between people without or with mild and moderate symptoms (supplementary table 4). The most common symptoms among participants with mild or moderate symptoms (n=75) were tiredness (60%), increased cough (61%), fever (47%), and headache (53%) (figure 2). Using Poisson regression, we found that participants with one or more comorbidities (IRR 1.93, 95%CI 1.40-2.64), and being infected during the period when the Delta variant was predominant (IRR 1.43, 95%CI 1.07-1.92) (Figure 3).

**Figure 2:**
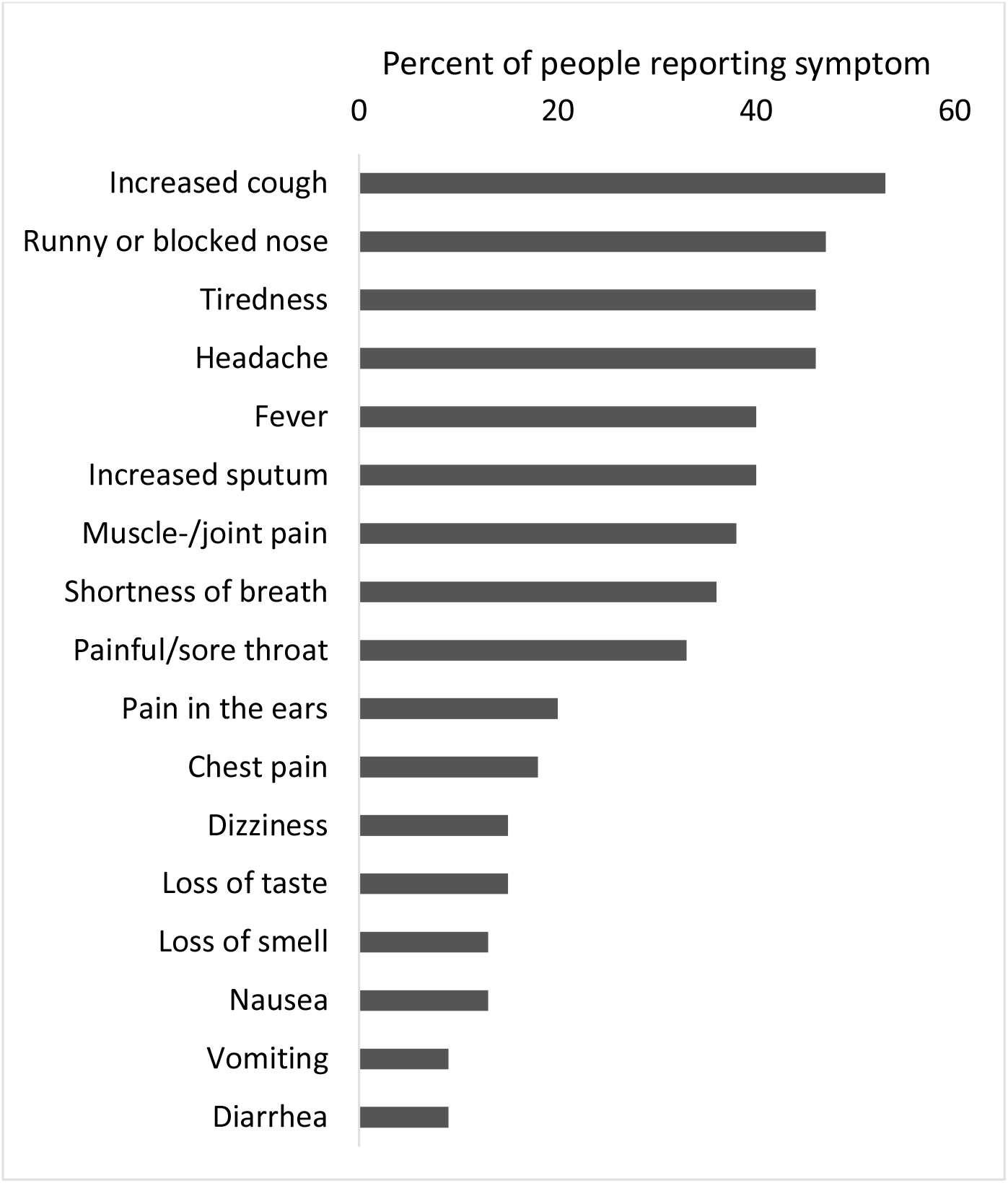
Reported symptoms among people with a SARS-CoV-2 infection (n=87), excluding re-infection.

**Figure 3:**
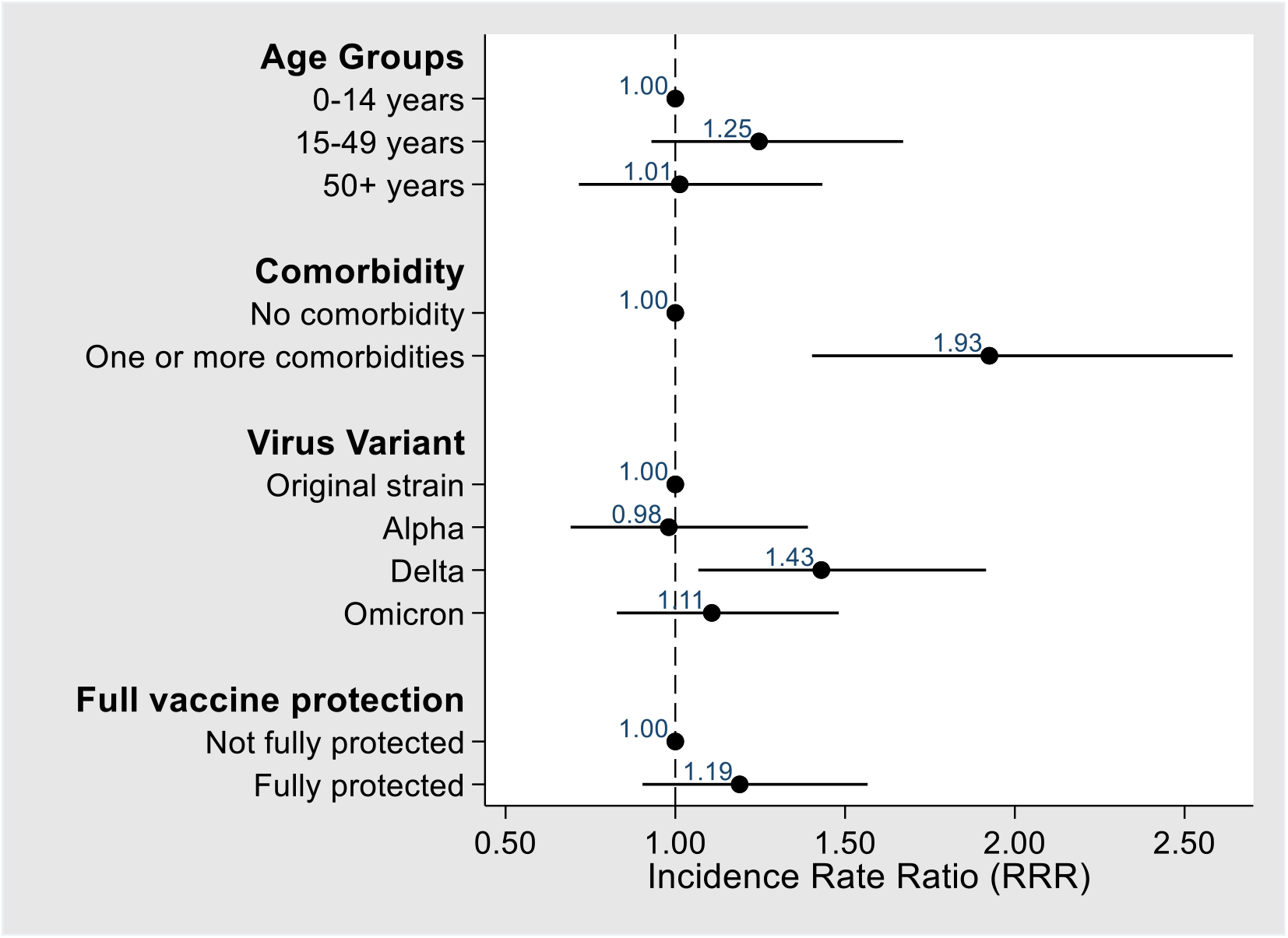
Factors associated with reporting more symptoms among COVID-PCD participants who reported a SARS-CoV-2 infection (N=87), excluding re-infections.

## Discussion

In this large participatory longitudinal study including more than 700 people with PCD, the cumulative incidence rate of reporting a SARS-CoV-2 infection was 9 per 100 PY. Children and people living in the UK were more likely to report a SARS-CoV-2 infection, and Delta and Omicron variants were also associated with higher likelihood of reporting an infection. Most participants reported no or mild symptoms; only 30% reported moderate symptoms. Four participants reported hospitalization, but none were treated in the intensive care unit or died. We found little difference in demographic and clinical characteristics between people reporting no, mild, and moderate symptoms. However, adults, participants with comorbidities, and people infected during the Delta wave were more likely to report multiple symptoms during a SARS-CoV-2 infection.

### Interpretation and clinical implications

The cumulative incidence of reporting a SARS-CoV-2 infection was 9 per 100 PY which was lower than what has been reported for the general population ^37^. Only Australia had a lower cumulative incidence rate in the general population than what we found among people with PCD (supplementary table 5). We observed the highest incidence rate in France (15 per 100 PY), but the cumulative incidence was also higher in the general population in France (19 per 100 PY) compared to other countries. Comparing incidence rate of reported SARS-CoV-2 in our study with whole-country incidence rates should however be done with caution due to differences in case definitions and methods for calculating follow-up time. The low incidence in our study population may be because people with PCD exhibited particular caution by reducing social contact, wearing masks in public, and getting vaccinated against COVID-19 ^18,23,24^. The extraordinary effort from PCD support groups taken to inform and encourage people with PCD throughout the pandemic may also have contributed to the low incidence. The incidence rate of 9 in the present study was higher than in our first publication from the COVID-PCD study using data collected until March 2021 where we reported an incidence rate of 3.2 per 100 PY ^18^. This is explained by the higher incidence rate during periods dominated by the Delta variant (IRR 4.5 in our study compared to the period with the original strain) and Omicron (IRR 13.8), and release of lockdowns and other strict public protection measures during the second year of the pandemic. The higher incidence rate during the Delta and Omicron period has also been shown for the general population ^29,38,39^. Omicron has increased resistance to antiviral immunity, which also affects the protective effect from vaccines ^40,41^.

Vaccination against COVID-19 was not associated with lower risk of reporting an infection, but this is probably due to the small number of non-vaccinated participants. Most adults in our study got vaccinated (95%) during the study period, and only one unvaccinated adult reported an infection after vaccination became available. Thus, after adjusting for age and calendar time, we had too little statistical power to show an effect.

Overall, severity of COVID-19 was mild in our study population with 14% of people reporting asymptomatic infection, 5% reporting hospitalization, and no observed deaths. Several factors may explain this. First, old age is one of the strongest predictors of severe COVID-19 ^42^, but only 8 people were older than 70 years in our population, which may be a reason why we observed no intensive care admissions or deaths. Secondly, 40% of the SARS-CoV-2 infections were reported during the Omicron-dominant period, which has been associated with less severe disease ^33,39^. Our results suggests that people with PCD may not be at higher risk of severe COVID-19 disease than people from the general population. However, although large for a rare disease, our study population is small compared to other studies, and it is difficult to assess generalizability of results from COVID-PCD participants to all people living with PCD. It is difficult to compare our results with studies in other chronic respiratory diseases, because no other similar longitudinal observational study exists.

We found no difference in demographic or clinical characteristics between people reporting no, mild, and moderate symptoms. This is probably because the severity of the reported cases was overall mild in our study. Even the four people reporting hospitalisation had mild or moderate symptoms. Other studies mainly described risk factors for intensive care treatment and mortality, outcomes which we did not observe. We did however find that age, presence of comorbidity, and virus variant were associated with reporting more symptoms among people reporting a SARS-CoV-2 infection, which is in line with risk factors identified in other studies ^32-34^.

### Strengths and limitations

A major strength of the study was the large study population for such a rare disease. The COVID-PCD is the largest study worldwide to collect data directly from people with PCD, and it represents all age groups and people with PCD from most places in the world. Another strength was the long follow-up starting in May 2020 and ending in May 2022. The weekly questionnaires minimized the risk that study participants would forget to report an infection with SARS-CoV-2. One reason for the high commitment of study participants to complete study questionnaires was that PCD support groups regularly encouraged members to participate in the study and keep completing questionnaires. A limitation of the study is that we only had self-reported data on test results for SARS-CoV-2. COVID-PCD is an observational study, and participants were not regularly tested for COVID-19 which may have led to an underestimation of the proportion of cases, due to unreported asymptomatic or mild cases. This would however not have affected the overall observed severity, as it is unlikely that a person with moderate or severe symptoms would not have been tested.

## Conclusion

While at the start of the pandemic, it was unclear if people with PCD were at high risk of severe COVID-19 disease, our results suggest that this is not the case. in this population of people with PCD, the incidence of reported SARS-CoV-2 was lower than in the general population and severity mainly mild. This may well be due to the particular caution that people with PCD took to avoid infection, careful use of facemasks, and early vaccination against COVID-19.

## Data Availability

All data produced in the present study are available upon reasonable request to the authors

## Declarations

### Ethics approval and consent to participate

The Bern Cantonal Ethics Committee (Kantonale Ethikkomission Bern) has approved this study (Study ID: 2020-00830). Informed consent to participate in the study is provided at registration into the study. Study participation is anonymous. Participants can withdraw their consent to participate at any time by contacting the study team.

### Availability of data and materials

The COVID-PCD data is available on reasonable request by contacting Claudia Kuehni by

### Competing interests

All authors declare no conflict of interest.

### Funding

This research was funded by the Swiss National Foundation (SNF 320030B_192804/1), Switzerland, the Swiss Lung Association, Switzerland (2021-08_Pedersen), and also received support from the PCD Foundation, United States; the Verein Kartagener Syndrom und Primäre Ciliäre Dyskinesie, Germany; the PCD Family Support Group, UK; and PCD Australia, Australia. M. Goutaki receives funding from the Swiss National Science Foundation (PZ00P3_185923). Study authors participate in the BEAT-PCD Clinical Research Collaboration, supported by the European Respiratory Society.

### Authors’ contributions

ESL Pedersen, LD Schreck, M Goutaki, and CE Kuehni made substantial contributions to the study concept and design. ESL Pedersen analysed the data and drafted the manuscript. ESL Pedersen, LD Schreck, M Goutaki, S Bellu, F Copeland, JS Lucas, M Zwahlen, and CE Kuehni critically revised and approved the manuscript.

## Acknowledgements

We thank all participants and their families, and we thank the PCD support groups and physicians who have advertised the study. We thank our collaborators who helped set up the COVID-PCD study: Cristina Ardura, Yin Ting Lam, Christina Mallet, Helena Koppe, Dominique Rubi, University of Bern, and Amanda Harris, University Hospital Southampton. We thank Francesca Santamaria, Federica Annunziata, and Vincenzo Miranda, Federico ll University, Naples, Italy, for helping to translate the study questionnaires to Italian.

---

**Supplementary table 1:**
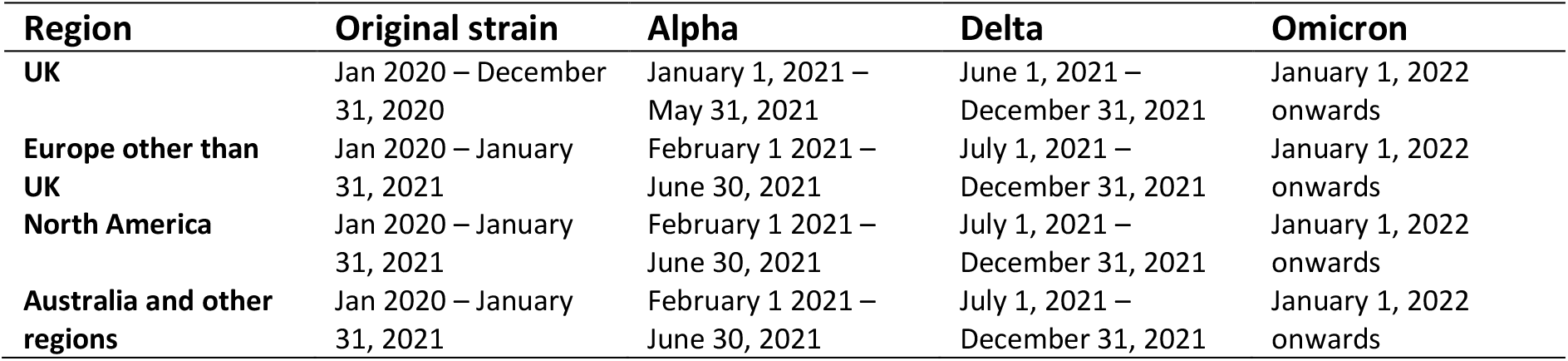
Definition of periods dominated by SARS-CoV-2 virus variants.

**Supplementary table 2:** Formulation of questions on SARS-CoV-2 infections, clinical characteristics, and co-morbidities in adults participating in COVID-PCD.

| Baseline | Question | Answer categories |
| --- | --- | --- |
| COVID-19 ever | Have you ever had COVID-19? | No; yes once; yes twice; yes three times, I don't know |
| Date of positive test | What date did you test positive the first time? (If you cannot remember the exact date, please select the date you think is correct) (asked for up to three time depending on the answer of question above) | date_dmy |
| Tested for COVID-19 | Have you been tested for COVID-19? | No; yes |
| Type of COVID-19 test | Which type of COVID-19 test was performed? | Viral test to identify a current COVID-19 infection usually done using a swab in the throat or nose; Antibody test to identify a past COVID-19 infection done by a blood test; Rapid antigen test to identify a current COVID-19 infection done using a swab in the throat or nose that gives a result within an hour; I don't know |
| COVID-19 test result | Was the test positive? | No; Yes; I don't know |
| COVID-19 test date | What date were you tested for COVID-19? (If you cannot remember the exact date, please select the date you think is correct) | date_dmy |
| Severity of symptoms | How seriously ill did you get during your COVID-19 infection? | No symptoms (asymptomatic); Mild symptoms (e.g. mild fever and/or cough); Severe symptoms (e.g. high fever, cough, headache, etc.) |
| Symptom | For this question, we would like you to think about symptoms that you experienced during your infection: Did you notice a worsening or new occurrence of the symptoms listed below during your COVID-19 infection? (Tick all that apply) | Fever/temperature; Chills; Muscle- /joint pain; Headache; Confusion; Increased cough; Increased sputum; Shortness of breath; Chest tightness; Chest pain; Sneezing; Runny/blocked nose; Loss of smell; Pressure or pain in the ears; Sore, scratchy, or painful throat; Difficulty swallowing; Loss of taste; Watery, red eyes; Tiredness or exhaustion; Loss of appetite; Dizziness; Nausea; Vomiting; Diarrhoea; Rash; Other symptoms |
| Hospitalization | Have you been in hospital (one night or more) because of COVID-19? | No; Yes |
| Hospitalization reason | Were you hospitalised because of COVID-19 or because of another reason? | I was hospitalised because of COVID-19; I was hospitalised for another reason (for example another infection or surgery), and the COVID-19 infection was detected with routine testing |
| Date of hospitalisation |  |  |
| COVID-19 vaccination | Have you been vaccinated against COVID-19? | Yes, I received one/two/three/four doses |
| Type of COVID-19 vaccine | Which vaccine did you get? | Pfizer-BioNTech (BNT162b2); Moderna (mRNA-1273); AstraZeneca, also called Oxford vaccine (AZD1222); Janssen/Johnson & Johnson (Ad26.COV2.S.); Sputnik V; CoronavAC (Sinovac); BBIBP-CorV (Sinopharm); EpiVacCorona; Convidicea (Ad5nCov) (CanSino Biologics); Covaxin (Bharat Biotech); Other; I don't know |
| Date of vaccination | What date did you get the first/second/third/fourth dose of vaccine? (If you cannot remember the exact date, please select the date you think is correct) | date_dmy |
| Bronchiectasis | Have you been diagnosed with bronchiectasis? (Bronchiectasis is a widening of the bronchi or airways caused by PCD, which can be seen in tests such as | No; Yes; I don't know |
|  | chest X-ray or CT (computer tomography) scans or MRI (magnetic resonance) examinations) |  |
| Lung function performed | Have you had your lung function measured in the past year? (By this we mean that you have blown through a tube and a computer has recorded your blow. This can be done in open space, or in a closed cabin) | No; Yes; I don't know |
| Result of FEV1 | What was the last FEV1? (This is the amount of air that can be blown out in 1 second; and is used to monitor lung function over time) | better than 90% predicted; 80-90% predicted; 70-80% predicted; 60-70% predicted; 50-60% predicted; below 40% predicted, I don't know |
| Asthma | Have you been diagnosed with asthma by a doctor and currently receive treatment? | No; Yes |
| Hypertension | Are you currently being treated for hypertension/high blood pressure? | No; Yes |
| Diabetes | Are you currently being treated for diabetes? | No; Yes |
| Heart disease | Are you currently being treated for heart disease or heart failure or have you had a myocardial infarction (heart attack) or coronary artery disease? | No; Yes |
| Cancer | Are you currently being treated for cancer? | No; Yes |
| IBD | Do you have inflammatory bowel disease (e.g. Crohn's disease or Colitis ulcerosa)? | No; Yes |
| Stroke | Have you ever had a stroke? | No; Yes |

**Supplementary table 3:** Proportion of infected participants and incidence rate (excluding re-infections) among people with primary ciliary dyskinesia in the COVID-PCD study (N=728) by age, sex, and country.

|  | N | Infected with SARS-CoV-2, n (%) | Incident infections (reported during follow-up) | Follow-up time (in person-years) | Incidence rate (infections per 100 person years, 95% CI) |
| --- | --- | --- | --- | --- | --- |
| **Total population** | 728 | 87 (12) | 62 | 716 | 9 (7-11) |
| **Age groups** |  |  |  |  |  |
| 0-14 years | 228 | 42 (18) | 35 | 218 | 16 (12-22) |
| 15-49 years | 381 | 32 (8) | 18 | 341 | 5 (3-8) |
| 50 years or more | 119 | 13 (11) | 9 | 157 | 6 (3-10) |
| **Sex** |  |  |  |  |  |
| Male | 292 | 39 (13) | 28 | 271 | 10 (7-15) |
| Female | 434 | 48 (11) | 34 | 443 | 8 (6-11) |
| **Country** |  |  |  |  |  |
| United Kingdom | 145 | 19 (13) | 19 | 159 | 12 (8-18) |
| USA | 128 | 13 (10) | 6 | 112 | 5 (3-12) |
| Germany | 103 | 18 (18) | 11 | 122 | 9 (5-16) |
| Switzerland | 48 | 2 (4) | 2 | 54 | 4 (1-14) |
| Italy | 47 | 3 (6) | 2 | 38 | 5 (1-20) |
| France | 42 | 6 (14) | 4 | 27 | 15 (6-37) |
| Australia | 32 | 3 (9) | 3 | 32 | 9 (3-28) |
| Other European countries | 123 | 16 (13) | 11 | 128 | 9 (5-15) |
| Other non-European countries | 59 | 6 (10) | 4 | 44 | 9 (3-23) |

**Supplementary table 4:** Associations between severity of COVID-19 disease and possible risk factors including demographic information, PCD disease severity, and vaccination status among all who reported a SARS-CoV-2 infection (N=87), excluding re-infections.

|  | **Asymptomatic**<br>**N=12** | **Mild**<br>**N=50** | **Moderate**<br>**N=25** | **p-value\*** |
| --- | --- | --- | --- | --- |
| **Age groups** |  |  |  | 0.829 |
| 0-14 years | 6 (14) | 26 (62) | 10 (24) |  |
| 15-49 years | 4 (13) | 18 (56) | 10 (31) |  |
| 50 year and older | 2 (15) | 6 (46) | 5 (38) |  |
| **Sex** |  |  |  | 0.623 |
| Male | 7 (18) | 22 (56) | 10 (26) |  |
| Female | 5 (10) | 28 (58) | 15 (31) |  |
| **Bronchiectasis** |  |  |  | 0.092 |
| No | 2 (6) | 25 (69) | 9 (25) |  |
| Yes | 10 (20) | 25 (49) | 16 (31) |  |
| **FEV1 (N=52)** |  |  |  | 0.855 |
| Above 60% predicted | 5 (16) | 16 (52) | 10 (32) |  |
| Below 60% predicted | 2 (10) | 12 (57) | 7 (33) |  |
| **Comorbidity** |  |  |  | 0.761 |
| No comorbidity | 12 (15) | 45 (57) | 22 (28) |  |
| One or more comorbidities | 0 | 5 (63) | 3 (38) |  |
| **Fully vaccinated at time of infection** |  |  |  | 0.489 |
| Not fully vaccinated | 7 (15) | 29 (62) | 11 (23) |  |
| Fully vaccinated | 5 (13) | 21 (53) | 14 (35) |  |

\*Fisher's exact

**Supplemenary table 5:** Proportion of infected people and cumulative incidence rate in the general population by 10.05.2022 for countries represented (by at least 10 persons) in COVID-PCD

| Country/area | Follow-up start date* | Follow-up years (in millions) | Population size | Total nr. cases | Cumulative IR per 100 person years |
| --- | --- | --- | --- | --- | --- |
| **COVID-PCD** | 30.05.2022 | 0.007 | 728 | 62 | 9 |
| **Australia** | 26.01.2020 | 58.9 | 25788217 | 6392018 | 10.9 |
| **Canada** | 23.01.2020 | 87.2 | 38067913 | 3814635 | 4.4 |
| **Denmark** | 02.02.2020 | 13.1 | 5813302 | 3124899 | 23.9 |
| **France** | 24.01.2020 | 154.3 | 67422000 | 29075412 | 18.8 |
| **Germany** | 27.01.2020 | 191.3 | 83900471 | 25503878 | 13.3 |
| **Ireland** | 29.02.2020 | 10.9 | 4982904 | 1527328 | 14.0 |
| **Italy** | 31.01.2020 | 137.0 | 60367471 | 16872618 | 12.3 |
| **Netherlands** | 27.02.2020 | 37.7 | 17173094 | 8073152 | 21.4 |
| **Norway** | 12.02.2020 | 12.2 | 5465629 | 1428699 | 11.7 |
| **Spain** | 01.02.2020 | 106.1 | 46745211 | 12058888 | 11.4 |
| **Switzerland** | 25.02.2020 | 19.2 | 8715494 | 3649155 | 19.0 |
| **USA** | 22.01.2020 | 763.8 | 332915074 | 82087703 | 10.7 |
| **United Kingdom** | 31.01.2020 | 154.8 | 68207114 | 22225565 | 14.4 |
| **Europe** | 23.01.2020 | 1716.3 | 748962983 | 195099900 | 11.4 |
| **North America** | 22.01.2020 | 1368.7 | 596581283 | 97151945 | 7.1 |

Data source: our world in data (https://ourworldindata.org/covid-cases, data downloaded on 11.06.2022).
\*Follow-up end date was set to 10.05.2022, same as in our study. Start date was defined as date of first case observed. Follow-up years was calculated as time between start date and end date times population size assuming that all persons were followed up the entire period of time.

## Notes

### Competing Interest Statement

The authors have declared no competing interest.

### Clinical Protocols

https://clinicaltrials.gov/ct2/show/NCT04602481?term=COVID-PCD&draw=2&rank=1

### Author Declarations

The ethics committee of the canton of Bern, Switzerland, approved the study (Study ID: 2020-00830).

